# HPV 51: A candidate for type-replacement following vaccination?

**DOI:** 10.1101/2021.07.27.21261121

**Authors:** Sarah J Bowden, Alison N Fiander, Samantha Hibbitts

## Abstract

**Background:** Human papilloma virus (HPV) infection is known to be a necessary cause of cervical cancer and is found in 99.7% of invasive cervical carcinomas. Current HPV vaccines protect against infection from strains 16 and 18. Baseline prevalence studies are important for the measurement of prophylactic vaccine impact and type-replacement monitoring. Between 2009-2010 the HPV research group in collaboration with Cervical Screening Wales conducted the ‘Base HPV 2009 study’ to determine baseline prevalence in unvaccinated women aged 20-22 in Wales. Preliminary analysis of results showed that in single HPV infection, 16 was the most prevalent high-risk strain followed by 18 and 51. This high prevalence of HPV 51 has not been observed in previous studies from Wales and is not a common finding elsewhere. This study aims to determine whether the high prevalence of HPV 51 observed in the Base HPV 2009 study is a true finding and if HPV 51 should be considered a candidate for type-replacement post-vaccination.

**Methods:** The first 100 single and 100 multiple HPV 51 positive liquid-based cytology (LBC) samples from the Base HPV 2009 study were selected for re-analysis. Each sample underwent DNA extraction and was tested using two methods: 1) Repeat of original methodology using GP5+/6+ HPV 51 PCR-ELISA. 2) HPV 51 E7 PCR. Data were then correlated with age, social deprivation score and cytology. 5 samples were excluded from analysis.

**Results:** Direct repeat of HPV 51 PCR-EIA identified 146 of 195 (75.0%) samples as HPV 51 positive. E7 PCR identified 166 of 195 (85.1%) samples as HPV 51 positive. When classified by cytological grade, the prevalence of confirmed HPV 51 increased with grade.

**Conclusions:** This study confirms that the prevalence of HPV 51 observed in the Base HPV 2009 study population is truly high and warrants further consideration. There is limited evidence on the cross-protection for 51 offered by the current HPV vaccine and it represents a potential candidate for type-replacement following vaccination. This study highlights the need for further longitudinal investigation into the regional and global prevalence of HPV 51. The data would recommend HPV 51 to be considered in future multivalent vaccines.

## Background

Human Papilloma Virus (HPV) infection is known to be a necessary and causal factor for the development of cervical cancer^1,2^ and is found in 99.7% of invasive cervical carcinomas^3,4^. HPV is highly prevalent and considered the most common sexually transmitted agent worldwide^2^. The lifetime chance of HPV infection is estimated to be 80%^5^. In the majority of women this infection is transient and will not result in cancer^3^. However, in a small number of cases, HPV infection persists, and invasive cervical carcinoma develops. Globally, cervical cancer is the 2^nd^ most common cancer in women and a leading cause of cancer mortality^2^.

Over 120 HPV strains have now been identified, of which 15-18 HPV types are known to be oncogenic and classified as high-risk (HR)^4-6^. HPV type-specific prevalence varies globally however types 16 and 18 are generally the most prevalent and are thought to be responsible for at least 70% of invasive cervical carcinomas^4,7^. These types therefore became the target of the first prophylactic HPV vaccines. Baseline study of the type-specific prevalence of HPV is important for assessment of the impact of prophylactic HPV vaccination and type-replacement monitoring.

Between April 2009-July 2010, the HPV Research Group at Cardiff University in collaboration with Cervical Screening Wales conducted the ‘Base HPV 2009’ study. 14,128 pseudo-anonymous LBC samples were collected from women aged 20-22 years, who had not been offered the HPV vaccine. Preliminary analysis of the Base HPV 2009 study showed that in single infections HPV 16 was the most prevalent (32.0%) followed by HPV 18 (11.2%) and HPV 51 (10.8%) (figure 1). HPV 51 prevalence among multiple infected samples was also high (18.4%). HPV genotype prevalence is heterogeneous worldwide. However, this high proportion of HPV 51 is not a common finding of previous prevalence studies^2^ and has not been seen in previous studies in Wales^3,8^.

**Figure 1:**
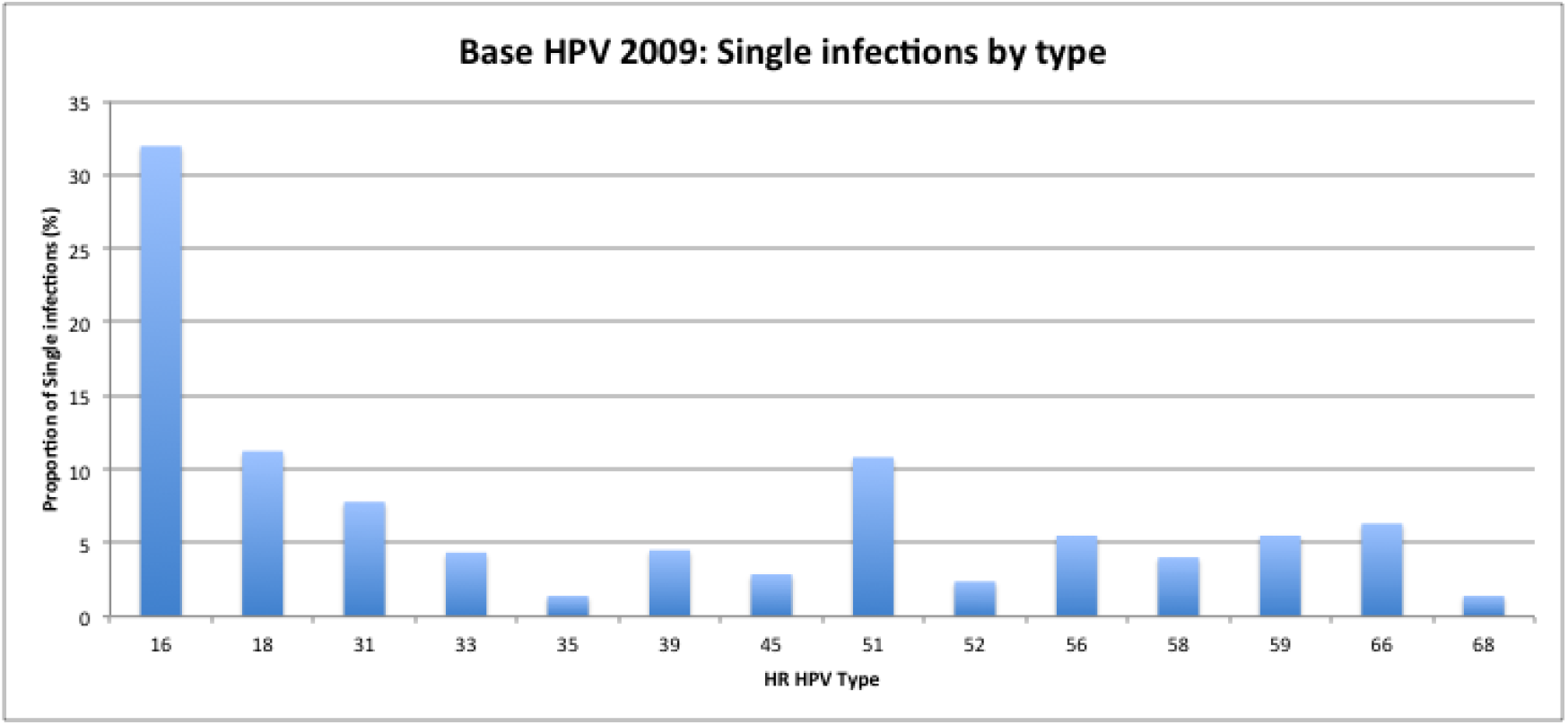
Proportion of single infections in the Base HPV 2009 study.

A high prevalence of HPV 51 was an unexpected finding of Base HPV 2009 and we have therefore chosen to re-analyse the HPV 51 results. Several issues may have led to false positives including contamination during sample processing and cross-hybridisation during HPV typing. Through retesting a selection of samples using two different methods, we aim to confirm the prevalence of HPV 51 observed and exclude the possibility of false positives.

In September 2008, the national HPV vaccination programme was implemented in the UK, offering Cervarix (an HPV-16/18 AS04-adjuvanted vaccine, GlaxoSmithKline Biologicals) to all girls between ages 12-13. Maximum efficacy is expected in women who have not been previously exposed to HPV^9^. It is hoped that vaccination will directly reduce the incidence of cervical cancer in vaccinated women as well as indirectly protecting unvaccinated women through the effects of herd immunity. Potential additional effects from vaccination include protection against other HPV-related cancers including anal, vaginal, vulval and oropharangeal^4^. In September 2012, the vaccine offered by the UK national programme was changed to Gardasil (quadrivalent vaccine active against HPV-16/18/6/11, Merck) so as to offer additional protection against low-risk HPV types responsible for genital warts.

One potential effect of vaccination is that a reduction in vaccine-targeted types may lead to type-replacement as the prevalence of other HR strains increase to fill the ecological gap^10,11^. Type-replacement has been observed in the past after introduction of the pneumococcal conjugate vaccine and *H.Influenza* type-b conjugate vaccine^12^. It is unknown whether type-replacement will occur following introduction of vaccination against HPV 16 and 18^11^. A high prevalence of HPV 51 could make it an important candidate for type-replacement. This has implications for future vaccine development and particularly any multivalent vaccine.

## Methods

### Specimen Collection

The Base HPV 2009 study involved the collection of 14,128 pseudo-anonymous LBC (BD SurePath, Source Bioscience) samples from unvaccinated women, aged 20-22 years. Samples were eligible if the woman was attending her first cervical smear and had an adequate cytology result available. The study was approved by Dyfed Powys Local Research Ethics Committee (08/WMW01/69).

LBC samples (Thinprep) were processed by the Cytology Laboratory according to the British Society of Clinical Cytology guidelines^13^. Residual cell pellets were re-suspended in the alcohol-based liquid and processed by the HPV Laboratory, Cardiff University.

### Specimen Selection

In the Base HPV 2009 study, 321 samples tested positive for HPV 51 (Figure 1). 132 samples were single infections of HPV 51 (singles) whilst 189 samples tested positive for HPV 51 and other HR HPV strains (multiples). In this study we chose to re-analyse 200 samples; the first 100 single and 100 multiple samples positive for HPV 51. The population characteristics for the selected samples are shown below (Table 1).

**Table 1:** Study population characteristics

|  | Single (n) | Multiple (n) |
| --- | --- | --- |
| **Age** |  |  |
| 20 | 62 | 65 |
| 21 | 23 | 18 |
| 22 | 15 | 17 |
| **Social deprivation score** |  |  |
| NA | 4 | 3 |
| Q1 | 17 | 20 |
| Q2 | 18 | 16 |
| Q3 | 23 | 17 |
| Q4 | 20 | 23 |
| Q5 | 18 | 21 |
| **Cytology** |  |  |
| Negative | 53 | 47 |
| Borderline | 22 | 26 |
| Mild | 23 | 22 |
| Moderate | 2 | 2 |
| Severe | 0 | 3 |

### DNA Extraction

DNA was extracted using the original methodology (appendix 1). A PCR for the human β-globin gene was performed on every DNA sample to determine extraction efficiency as described earlier^8^.

### HPV Testing

All samples were re-analysed using the following methodology:

1. **Repeat of original GP5+/6+ ELISA:** The GP5+/6+ HPV PCR-ELISA method of Jacobs *et al*^14^ was performed on all specimens in a 96-well format with minor modifications (appendix 2).
2. **HPV 51 type-specific E7 Linear PCR followed by E7 Nested PCR (appendices 3 and 4**)
3. **Repeat of Discordant Results:** Where there was discordance between GP5+/6+ ELISA and E7 PCR results, samples were rested using both methods. During the course of this project, a PhD student saw improved specificity with the use of Hotstar Taq instead of Invitrogen Taq in E7 PCR (figures 2 and 3). We therefore used Hotstar Taq when repeating discordants (appendix 5).
4. **Repeat of all samples with E7 PCR using Hotstar Taq:** Repeat of discordant samples with the Hotstar Taq method (appendix 5) showed an increased specificity of the assay (figures 2 and 3). We therefore decided it was appropriate to repeat all samples using the Hotstar Taq method.
5. **Sequencing:** To confirm the specificity of the E7 PCR method we sent a selection of positive samples for sequencing.

### Statistical Analysis

Statistical analysis was performed on all samples that were β-globin or HPV 51 positive. HPV 51 GP5+/6+ ELISA positive samples were defined as samples that tested positive in the original Base HPV 2009 study and also positive in either the 2nd or 3rd repeat ELISA i.e. at least 2 of 3 positive results. HPV 51 E7 PCR positive samples were defined as samples that tested positive in the E7 PCR test of all samples using the Hotstar Taq method (as described in stage 4 above).

**Figure 2.**
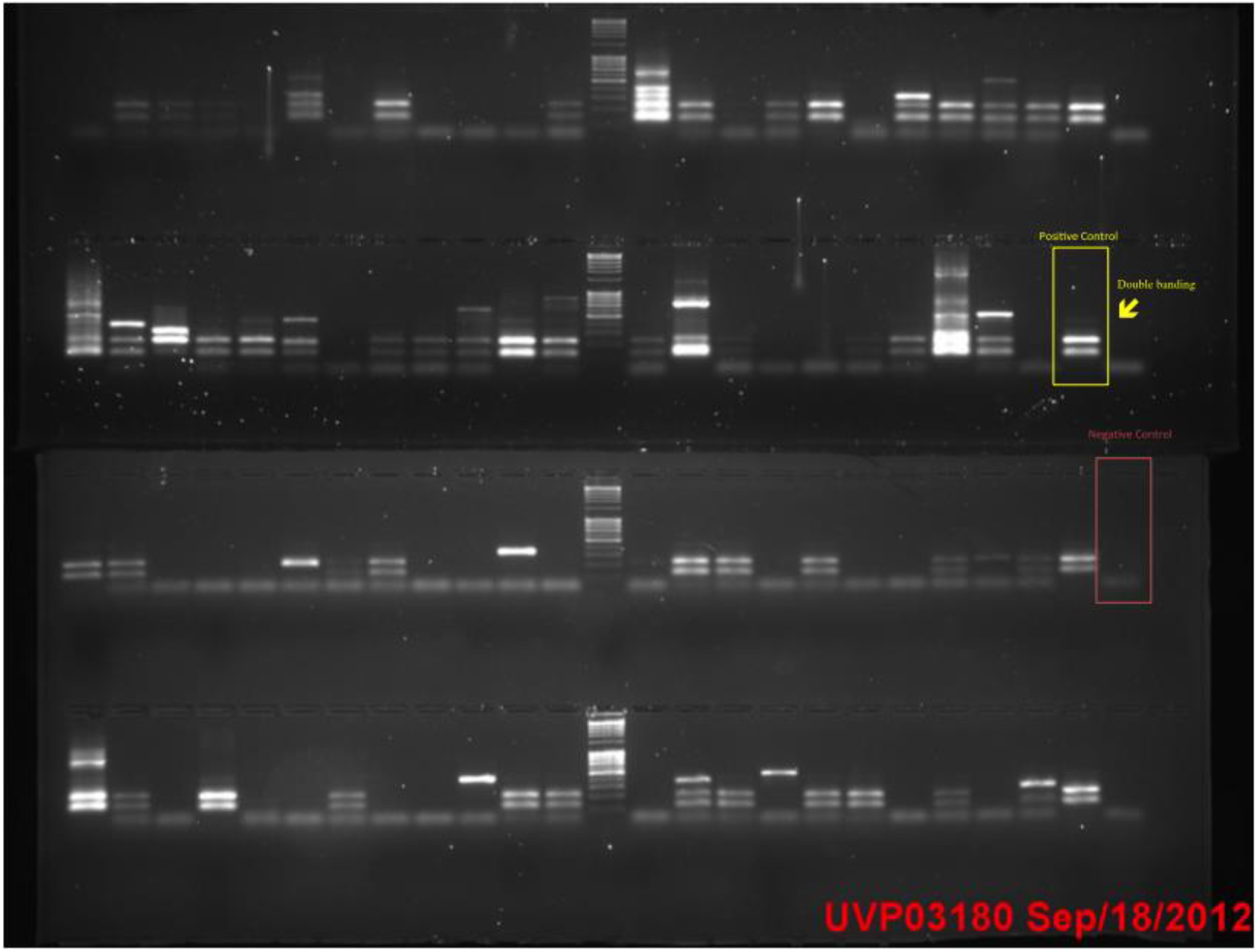
E7 PCR using Invitrogen Taq. Yellow box highlights the double banding in positive samples showing an unexpected fragment.

**Figure 3:**
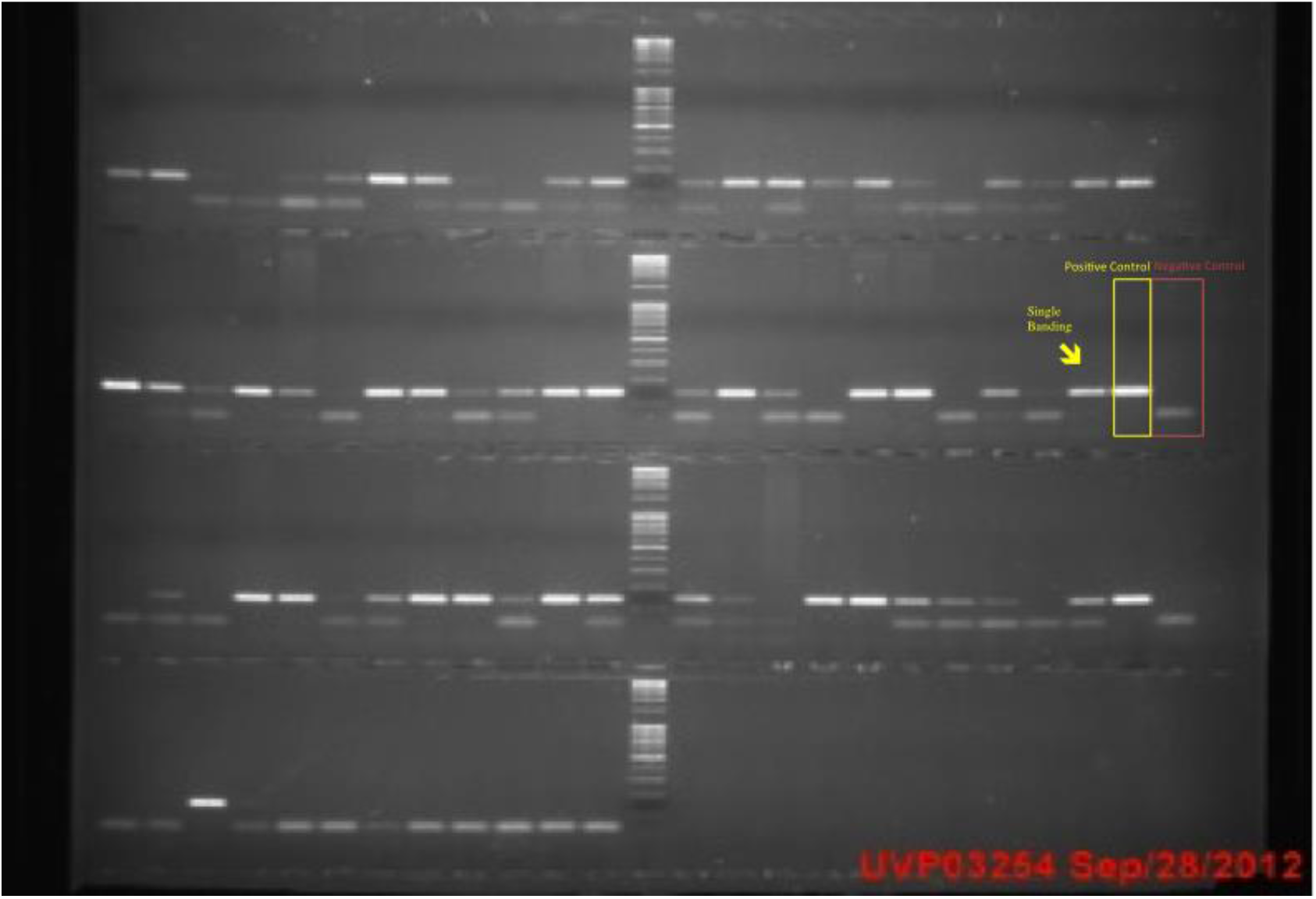
E7 PCR using Hotstar Taq. Yellow box highlights the single band of expected fragment only.

Data on cytology, age and social deprivation status for each sample were aligned with repeated HPV test results using GP5+/6+ and E7 methods. Social deprivation was estimated in the original Base HPV 2009 Study by linking postcodes to the Welsh Index of Multiple Deprivation which describes levels of deprivation across Wales with higher scores indicating greater deprivation^15^.

Association between tests was calculated using Kappa analysis. Fisher’s exact test was used to calculate p-values where appropriate.

## Results

### Sample Adequacy

5 samples were found to be β-globin PCR and HPV 51 negative and were deemed inadequate and excluded from statistical analysis (figure 4). 1 of these samples was single infection, 4 multiple. Therefore 99 singles and 96 multiples totaling 195 samples were included for analysis.

**Figure 4:**
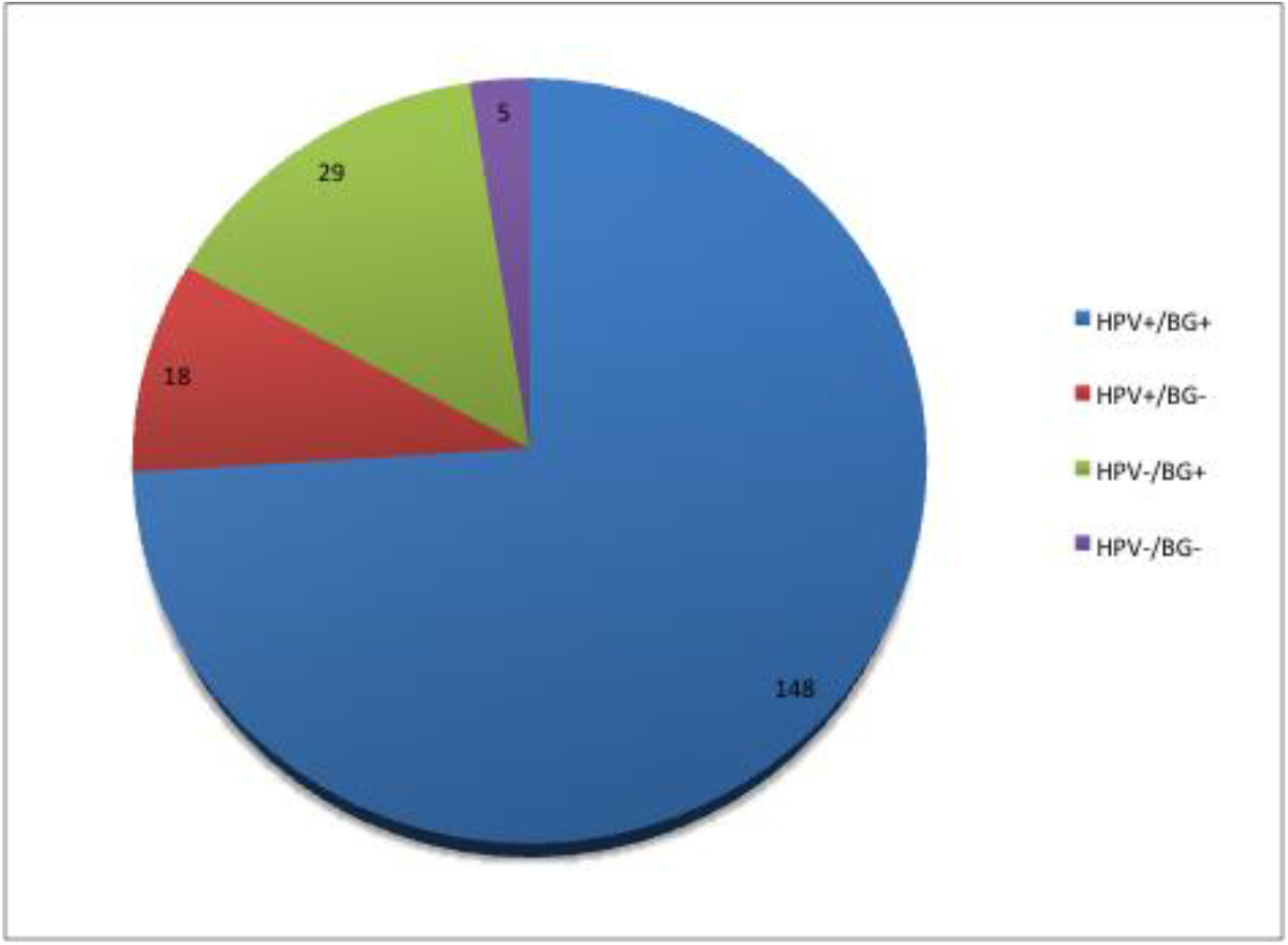
Comparison of HPV 51+ and β-globin+ results

### GP5+/6+ PCR ELISA

Re-analysis using GP5+/6+ PCR ELISA found that 146 of 195 samples (75.0%) tested HPV 51 positive. Of these 78 singles and 68 of multiples retested HPV 51+ positive (figure 5).

**Figure 5.**
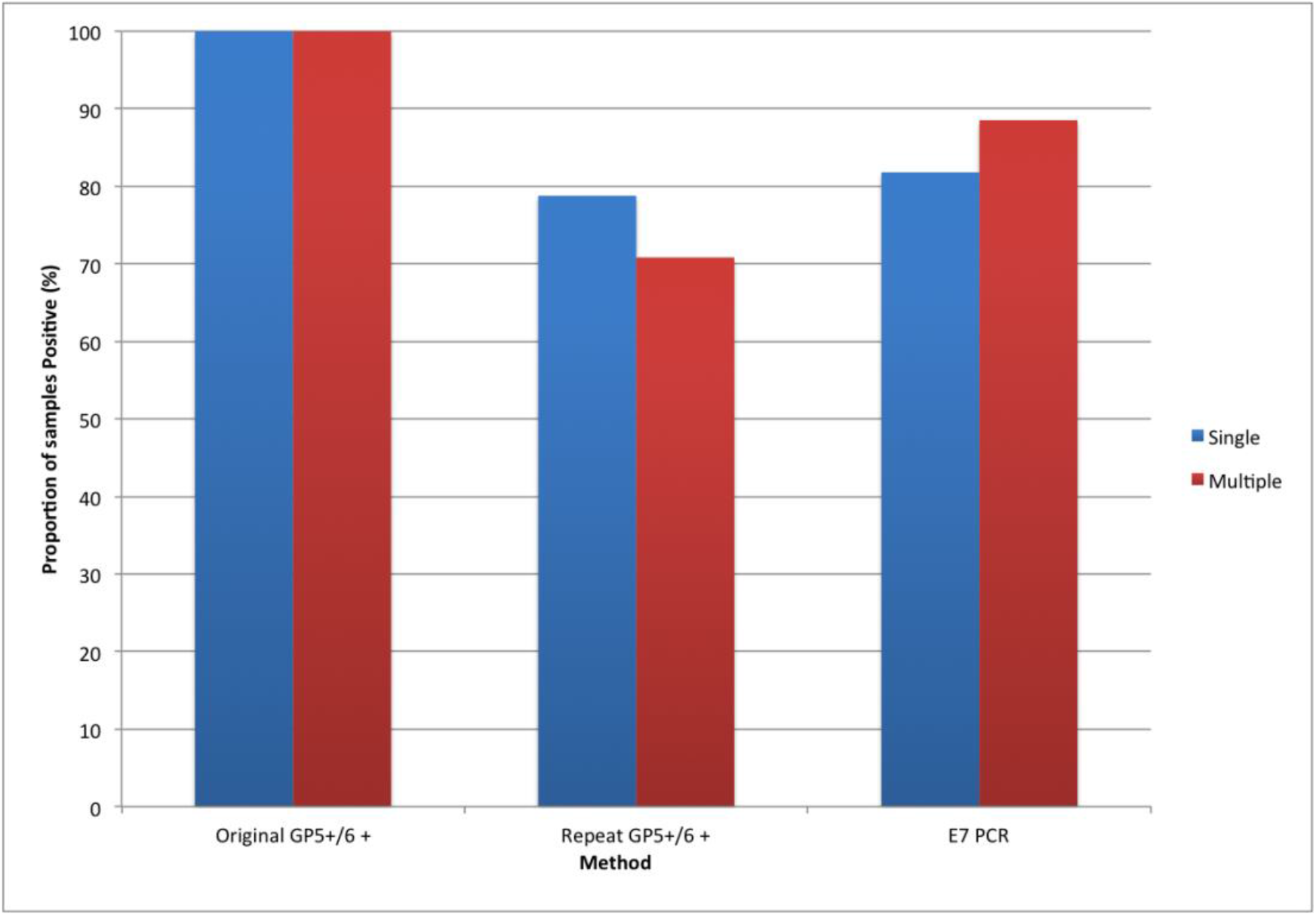
Proportion of samples testing HPV 51 positive by test: singles and multiples.

### E7 PCR

On re-analysis of the samples using E7 HPV 51 PCR it was found that 166/195 (85.1%) of samples tested positive. 81 were singles and 85 were multiples (figure 5).

5 samples tested positive by repeat GP5+/6+ ELISA and negative by E7 PCR. 25 samples tested positive by E7 PCR and negative by repeat GP5+/6+ ELISA (figure 6). Kappa analysis showed there was a moderate agreement between the repeat GP5+/6+ ELISA and E7 PCR tests (κ=0.527 (95% CI 0.384 to 0.670)).

**Figure 6.**
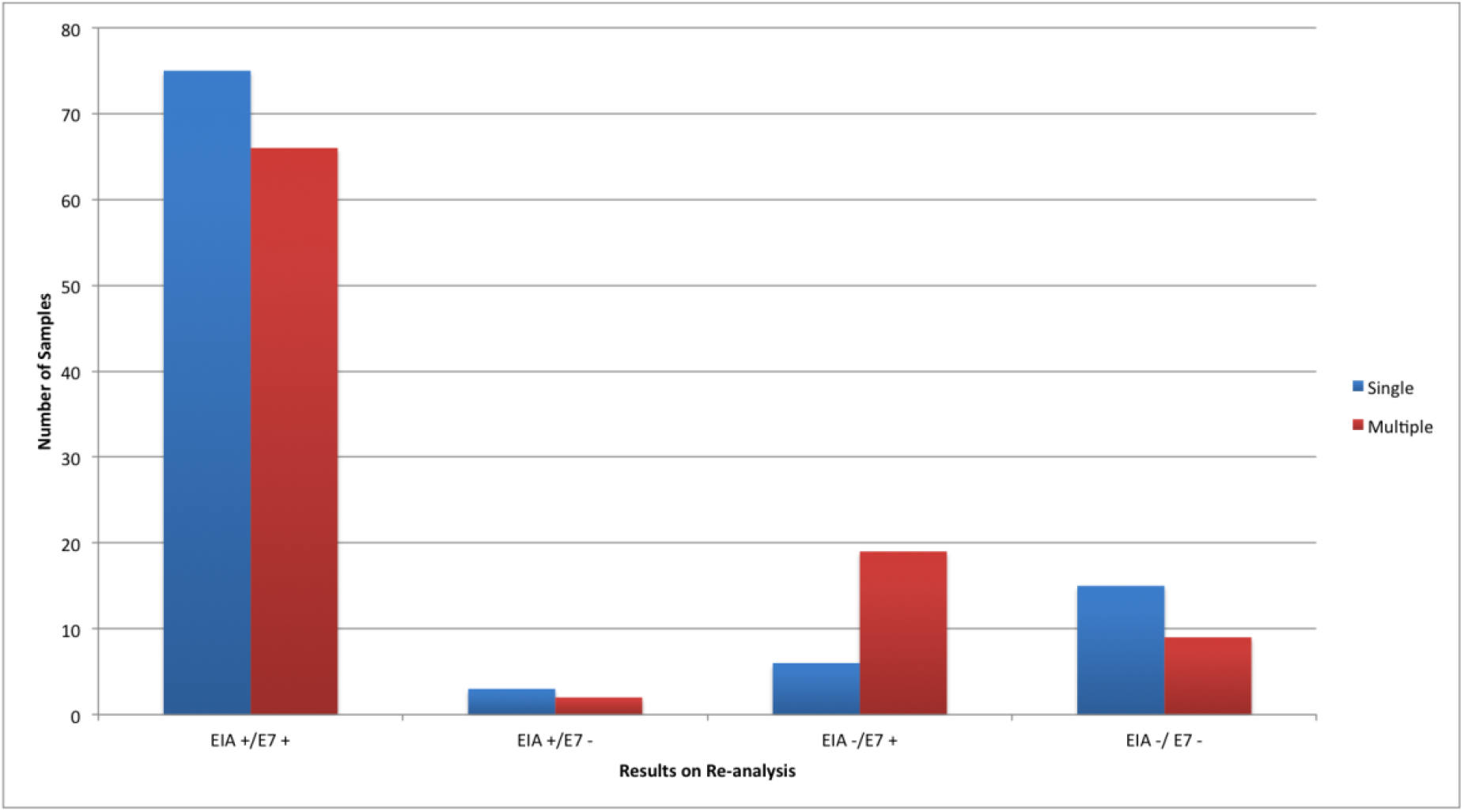
HPV 51 results from re-analysis with GP5+/6+ ELISA (EIA) and E7 PCR (E7): singles and multiples.

### Stratification of results

Results were stratified by cytology, age and SDS. HPV 51 negative samples on repeat GP5+/6+ ELISA, were more commonly of a low cytological grade (figure 7). The number of HPV 51 negative samples decreased as grade increased. No moderate or severe cytology samples retested negative for HPV 51 using E7 PCR. One severe but no moderate samples retested negative for HPV 51 using GP5+/6+.

**Figure 7.**
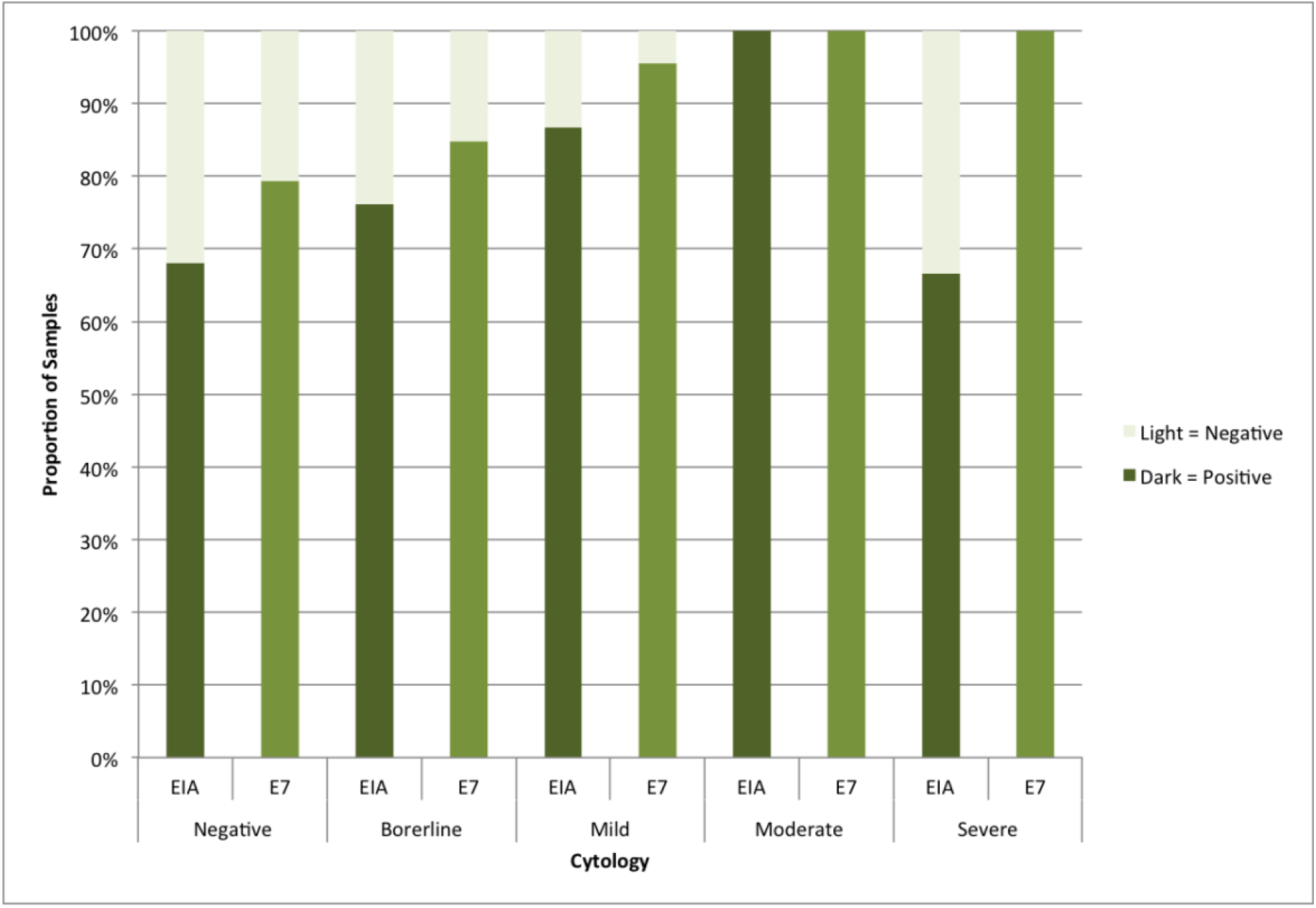
Proportion of samples positive stratified by cytological grade

There was no observed difference between results stratified by age or SDS (figures 8 and 9).

**Figure 8.**
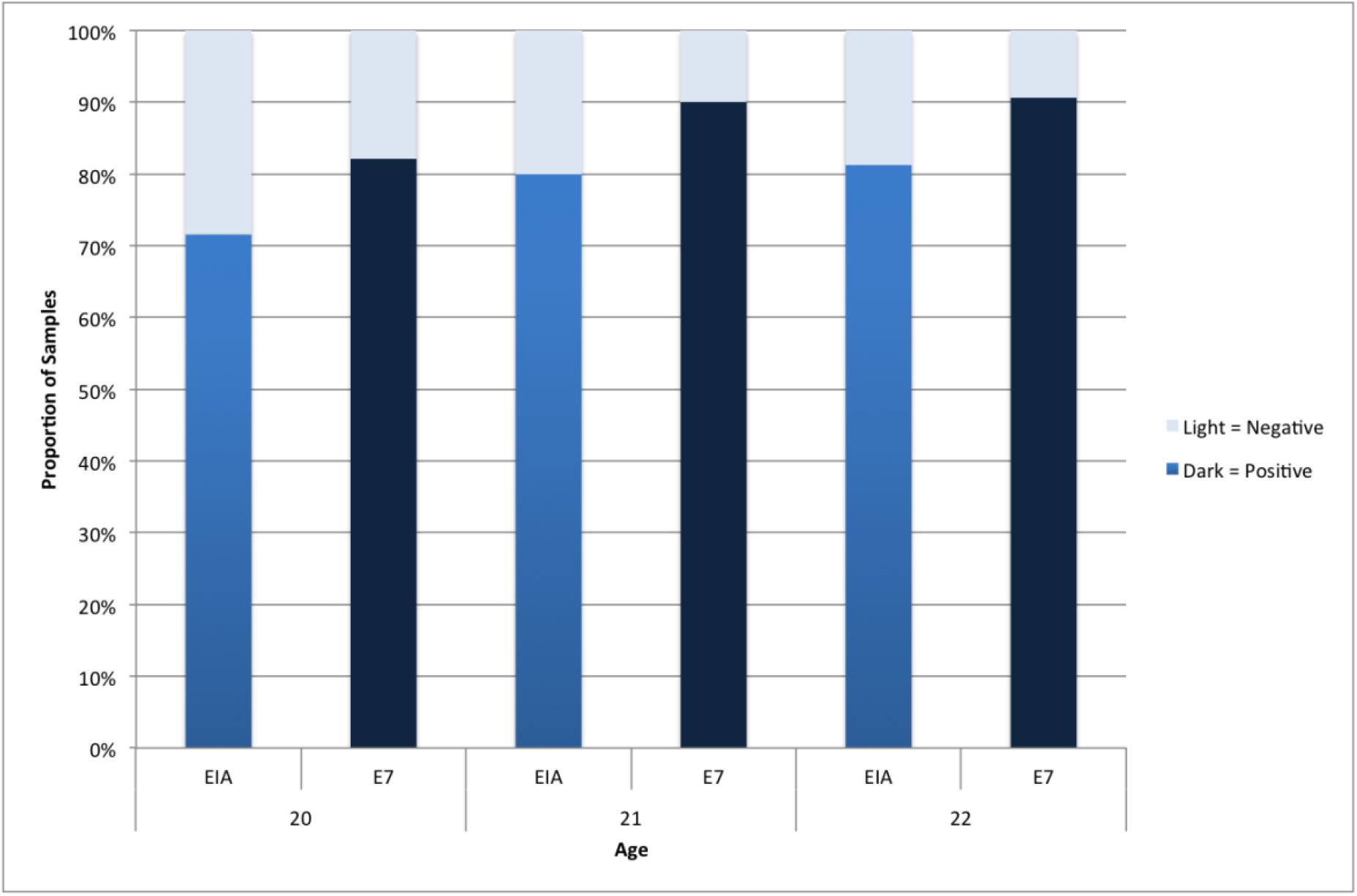
Proportion of samples positive stratified by Age

**Figure 9.**
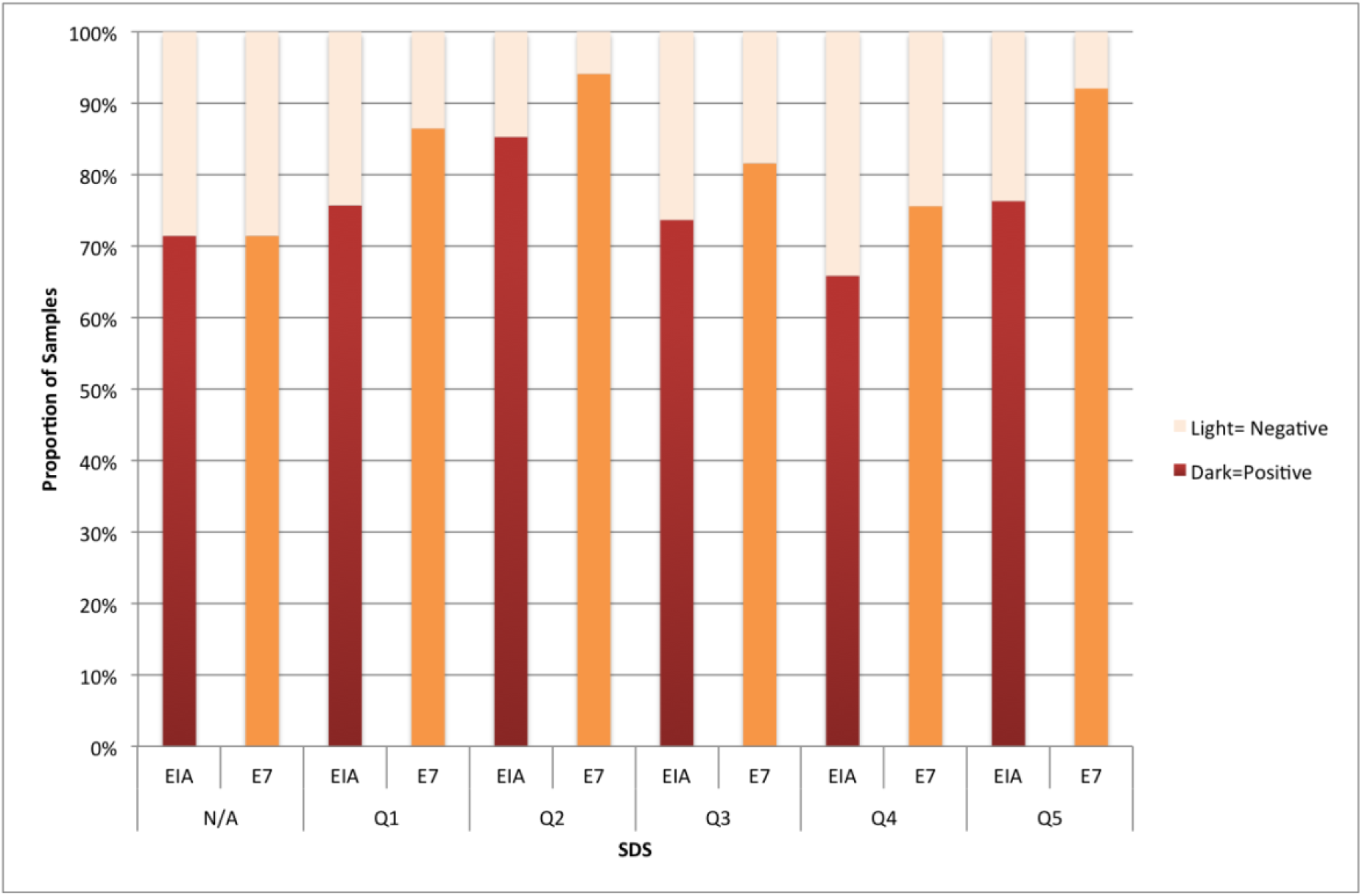
Proportion of samples positive stratified by social deprivation score.

### Multiple Infection Samples

We compared the type prevalence seen in all multiples with the type-prevalence seen in multiples which tested negative on reanalysis by GP5+/6+ ELISA or E7 PCR (figure 10). The proportion of HPV 16 appeared to be higher in samples retesting negative by E7 PCR compared to GP5+/6+ negatives and all multiples. However this was not found to be statistically significant (p=0.4094 and p=0.3016 respectively).

**Figure 10.**
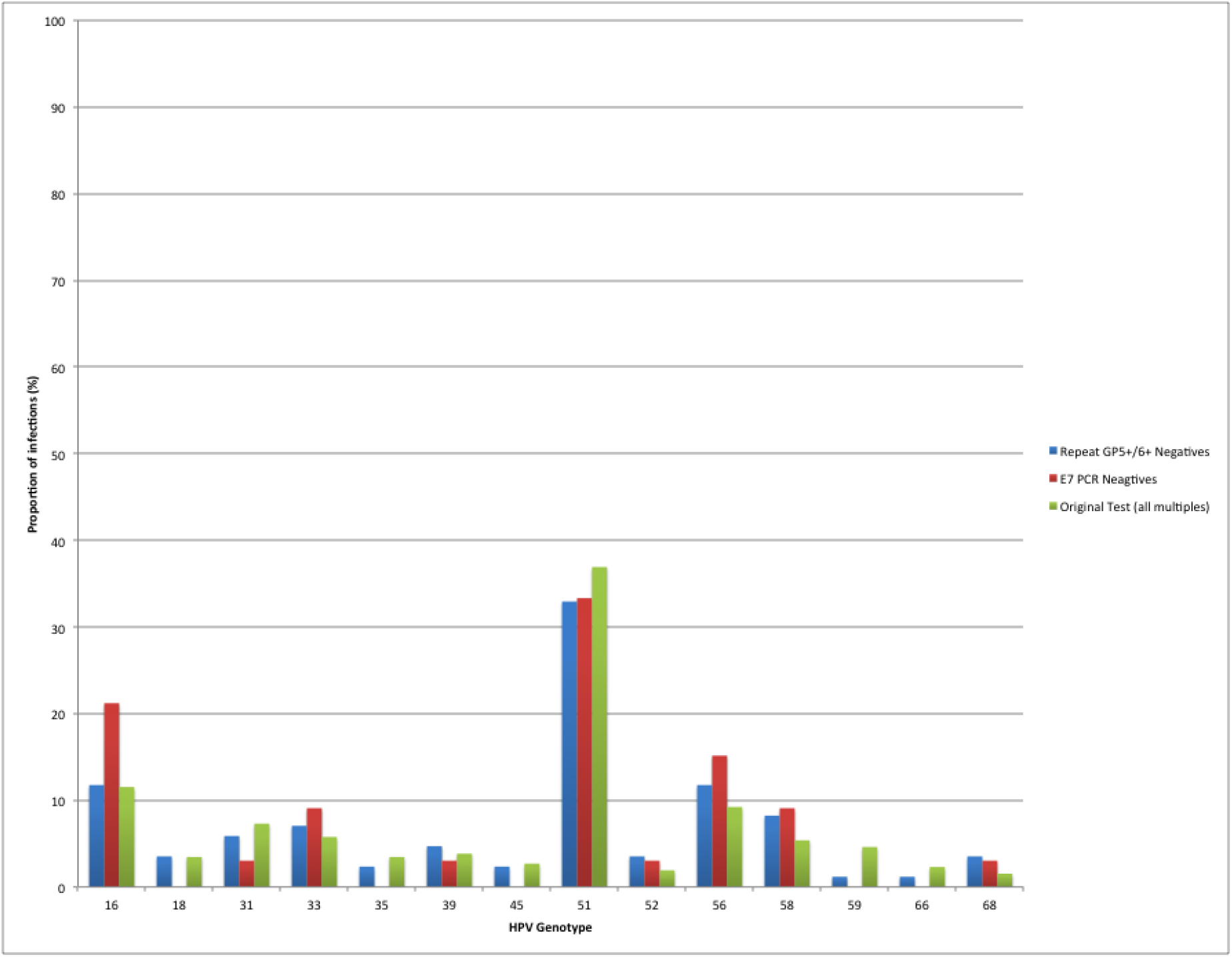
Proportion of infections in multiple samples. Typing results from Base HPV 2009 used to compare all multiples, GP5+/6+ repeat negative multiples and E7 negative multiples.

### Sequencing

Seven samples were sent for sequencing to confirm the specificity of our E7 51 PCR assay (figure 11). Analysis was undertaken using 4 peaks® software and a BLAST® search using megablast. 5 sequences were found to be valid which aligned with two patients. Analysis showed there was no alignment with any known human sequence. Four known alignments were seen for each of the 5 samples sent: the closest alignment was a 99% match to the known HPV 51 E7 region and 3 alignments of an 85-89% match to HPV 82 E7 region were also observed.

**Figure 11.**
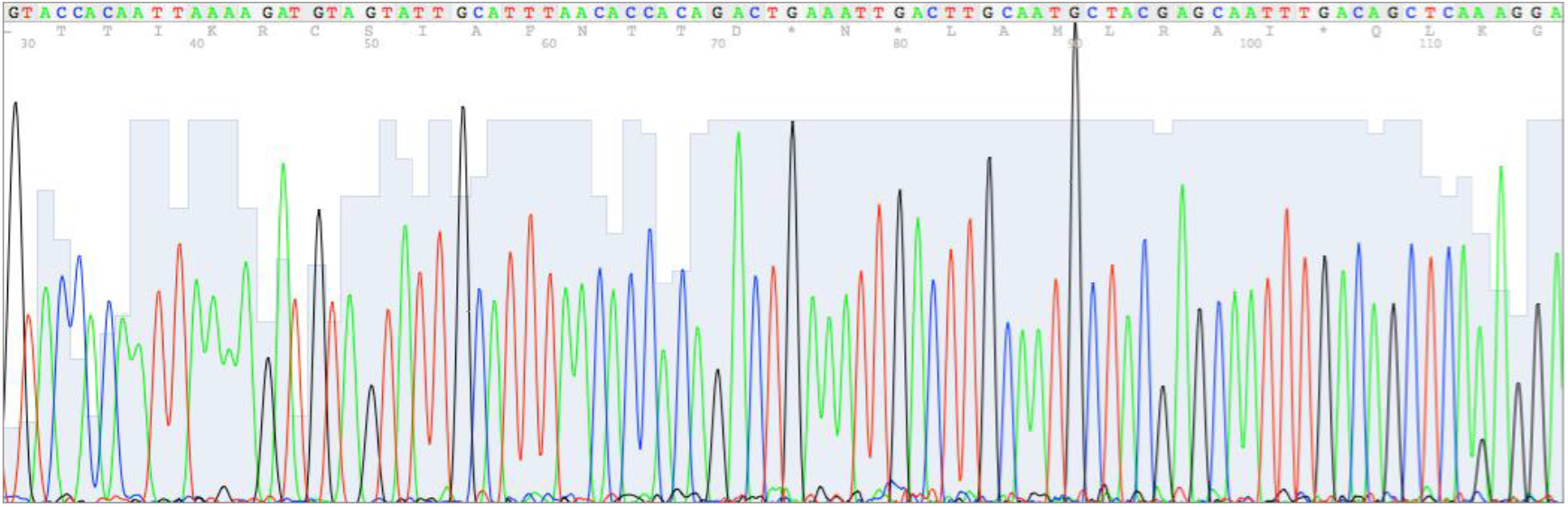
Example of sequencing data for one sample; shows a 99% match to known HPV 51 E7 region.

## Discussion

The high prevalence of HPV 51 observed by the Base HPV 2009 study raised the question of whether false positive results had arisen. An initial hypothesis for false positives was that cross-hybridisation between the HPV51 probe and other HPV types could have occurred during genotyping. To test this hypothesis we used E7 type-specific PCR specific to HPV 51. If cross-hybridisation had occurred in the original test we would expect less multiple compared to single infection samples to retest positive by E7 PCR. We observed no significant difference between single and multiple infections therefore making cross-hybridisation unlikely (figure 4). Additionally we found that multiples were more likely than singles to test negative by repeat GP5+/6+ PCR ELISA but positive by the more specific E7 PCR method (figure 3); the positivity of multiples in the more specific method supports the above assumption that cross-hybridisation did not occur.

Comparison of multiple infection samples suggested that type 16 was of higher proportion in samples that tested negative by E7 51 PCR compared to all multiples. However this was not found to be significant and therefore there is no evidence to support cross-hybridisation with HPV 16, or any other tested type, in the Base HPV 2009 GP5+/6+ assay.

To confirm the specificity of our E7 PCR method we sent a selection of positive samples for sequencing (figure 11). Sequencing showed a 99% match to the known HPV 51 E7 sequence in all 5 samples. There was also a close match (85-89%) to HPV 82, which is a low prevalence HR HPV known to be phylogentically similar and of the same A5 species as HPV 51^16,17^ (figure 12). Type 82 is not included in our current type-specific assay and therefore the prevalence in the Base HPV 2009 population is unknown. However the genetic sequences of 82 and 51 are sufficiently different to make the chance of HPV 82 being detected through E7 51 PCR unlikely.

**Figure 12.**
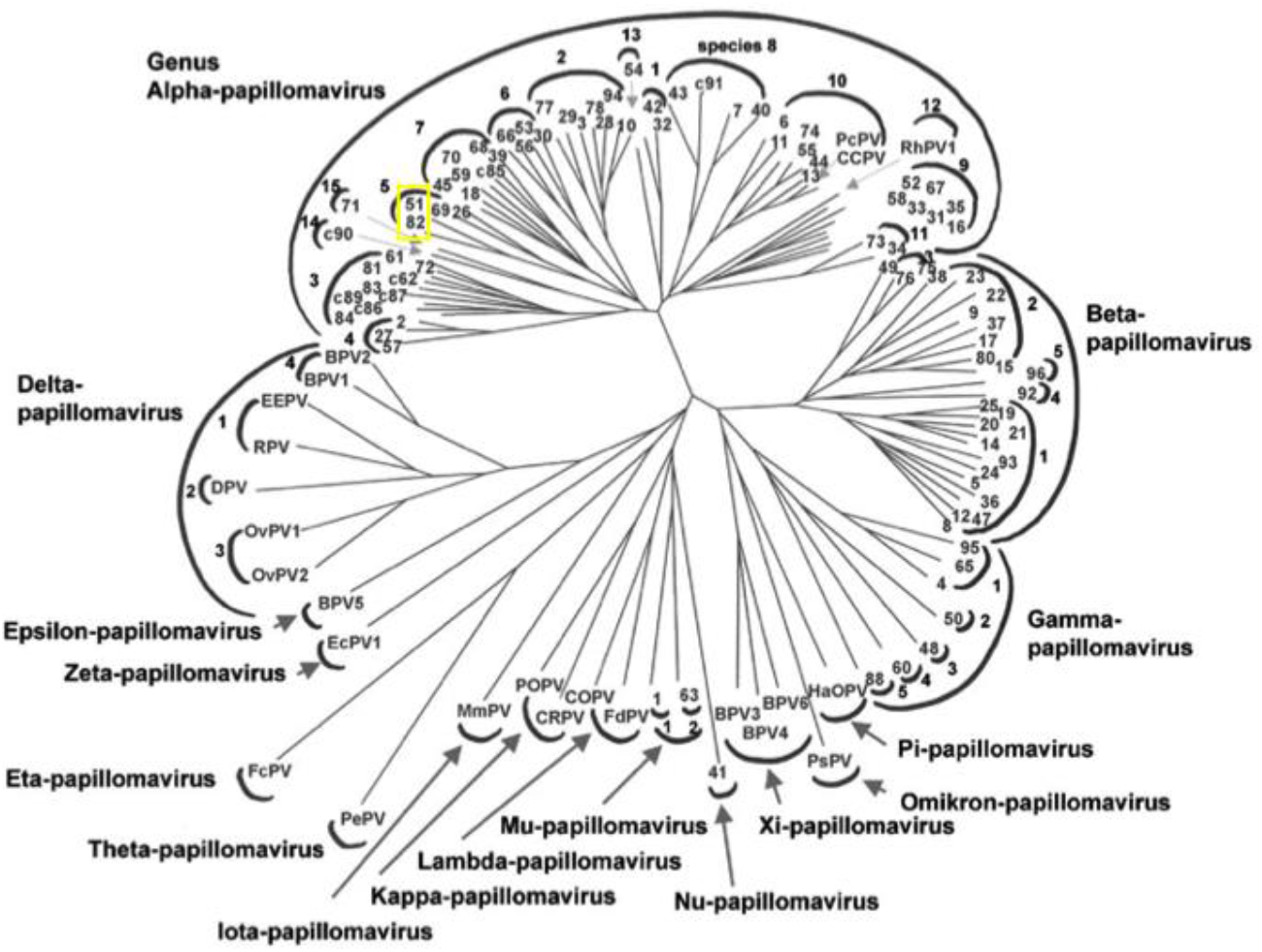
Phylogenetics of HPV including HPV 51 in the A5 species. *Adapted from de Villiers et al*^17^

Our results showed that samples of high-grade cytology were most likely to retest positive and samples of negative cytology were most likely to retest negative. We would expect dyskaryotic samples to be HPV positive and therefore this supports the validity of our re-analysis. There was no significant association seen for age or SDS.

Reanalysis confirms that at least 85.1% of Base HPV 2009, HPV 51 positive samples are true 51 positives (figure 5). 14.9% of samples therefore did not test positive on re-analysis and could have been false positive on original testing. Contamination is a significant problem in molecular biology and may explain false positives. Contamination could have occurred during original testing, in probe synthesis or during setup of the HPV 51 plasmid. Viral aerosol in the laboratory could also have led to contamination. Variation in setup may§ have contributed to false positive or negative results; the reproducibility of GP5+/6+ has previously been found to be around 86%^18^. Where resources are not a limiting factor, every test should be repeated in triplicate to ensure reproducibility.

Our results have confirmed that a high HPV 51 prevalence exists in unvaccinated women aged 20-22 in Wales. A high proportion of HPV 51 has recently been seen in several prevalence studies undertaken in Scotland^19,20^[personal communication, Kate Cuschieri], Italy^21^ and small studies of both HIV positive^22^ and negative women in Sao Paulo, Brazil^23^. However in a global meta-analysis undertaken in 2007, HPV 51 did not emerge as a highly prevalent genotype^2^. This raises the questions of whether HPV 51 is a significant genotype in other unknown areas and whether its prevalence has increased since previous studies.

The significance of a high prevalence of HPV 51 for cervical cancer depends on several factors including the carcinogenicity of type 51. HPV 51 has been classified as definitely carcinogenic since 2005 but is thought to be less carcinogenic than HPV 16 and 18^24,25^. HPV 51 is currently thought to account for around 1.16% of cervical cancer^24^. A recent study of invasive cancers in Wales, found no cancers to be positive for HPV 51^7^. HPV 51 is therefore a definite carcinogen but may currently account for a low proportion of invasive cancers.

The efficacy of prophylactic HPV vaccination is related to the proportion of cancers attributable to vaccine-types and the potential for HPV vaccines to confer cross-immunity for cancers caused by non-vaccine types^4^. HPV 51 belongs to the A5 species, which is phylogenetically different to the A9 species of HPV 16 and 18. One recent study observed that some cross-protection exists for 51 following vaccination^1^. However, others suggest that current licensed vaccines provide limited cross-immunity and protection against phylogenetically-different but clinically important HR HPV types^1,6,26-28^. Cancer caused by HPV 51 may not be protected against under current vaccination schemes. This has important consequences where HPV 51 is highly prevalent.

It is unknown whether type-replacement will occur in the long-term following prophylactic vaccination. The likelihood of type-replacement is dependent on whether HPV types are synergistic, unrelated or competitive in their infection of the host and also on the cross-immunity of the vaccine used^10,12^. If HPV types are synergistic then any vaccine which decreases the prevalence of vaccine-types will lead to a reduction in infections by other types as has been suggested by several studies and mathematical models^26,27,29^. *Chaturvedi et al*^30^ found that infection by different HPV types is unrelated and therefore that type-replacement is unlikely to occur. However some suggest that HPV may compete for host infection and that a reduction in vaccine-types may lead to an ecological niche for the increase of non-vaccine types^31^. *Durham et al* believe that epidemiological data supports cross-immunity between HPV types which in turn may cause significant type-replacement following vaccination^12^. In summary, continued monitoring of population specific and type specific prevalence of HPV is necessary to draw conclusions on the impact of prophylactic vaccination and occurrence of type-replacement. If type-replacement occurs then HPV 51 may account for a greater number of cervical cancers than present.

*Tumban et al*^6^ have suggested that the current vaccines based on virus like particles of the L1 protein are highly type-specific and therefore have little effect on phylogenetically different types. L2-based vaccines are currently under investigation and are thought to confer increased cross-immunity. However currently L2 vaccines elicit lower neutralizing antibody titres than L1 vaccines. If this issue can be addressed then future L2 vaccines may allow protection against a wider range of HPV types allowing improved efficacy against cancer caused by non-vaccine types.

It has been suggested that a multivalent vaccine including the 8 most prevalent HPV types could increase protection to >90% of cervical cancers^11^. However, prevalence of HPV types varies greatly worldwide. Further investigation into the prevalence of HPV 51 is necessary to determine the need for its inclusion in any future multivalent vaccine.

HPV testing for triage was introduced into the NHS Cervical Screening Programme for England in April 2011^32^. There is ongoing research into the role of HPV testing for use in clinical practice. A high prevalence of HPV 51, as observed in the Base HPV 2009 study, warrants the recommendation that type 51 is included in any type-specific HPV test used in clinical practice.

### Limitations

The results of this study were confined to the limitations of the assay. We saw variability in intra-assay and inter-assay results, which could have been misleading.

Due to resource and time constraints not all samples that tested positive for HPV 51 in the Base HPV 2009 study were retested. 200 of 321 samples were selected, more samples could have been retested which would have led to a larger sample size and better estimate of the proportion of samples which were truly HPV positive.

Five samples were excluded as inadequate as defined by both HPV 51 and β-globin negativity. These samples could have differed from samples included. If more resources are available these samples should be re-extracted and retested.

## Conclusions

There is inconclusive evidence on whether type-replacement will occur following prophylactic HPV vaccination. Continued monitoring of type-specific prevalence in necessary to determine whether type-replacement is occurring and the impact of vaccination on HPV prevalence.

Our study confirms a high prevalence of HPV 51 in unvaccinated women aged 20-22 in Wales, high prevalence has also been seen in women of varying ages in Scotland, Italy and Sao Paulo, Brazil. HPV 51 is phylogenetically distinct from HPV 16 and 18 and therefore unlikely to be cross-neutralised by current L1-based HPV-16/18 vaccines. HPV 51 could therefore continue to cause invasive cervical cancer and in the event of type-replacement, cause an increased proportion of invasive cervical cancer in the future. Findings from the study [9] would recommend that HPV 51 be included in UK HPV screening programmes and testing of CIN and cancerous lesions to ensure continued detection and monitoring.

HPV vaccination is potentially a major public health breakthrough, however it is essential that research into the population-based impact of widespread vaccination continues to determine efficacy and monitor epidemiological change. Further investigation into the prevalence of HPV 51 and its carcinogenicity is important to inform the needs of future HPV vaccination and HPV testing in clinical practice.

## Data Availability

Data will be made available on reasonable request

## Abbreviations

HPV: human papillomavirus
CIN: cervical intraepithelial neoplasia

## Conflict of Interest Declaration

### Ethical approval

Not required

## Acknowledgements

We gratefully acknowledge all members of the HPV Research Team at Cardiff University for their contributions, in particular Angharad Edwards and Rachel Houghton. Authors’ contributions are: Sarah Bowden (Corresponding author): data acquisition, analysis and interpretation, drafting of manuscript and final approval of manuscript; Alison Fiander: conception, critical review of the paper and final approval of manuscript; Samantha Hibbitts: conception, project design and management, data acquisition and analysis and interpretation, drafting and critical review of the manuscript, final approval of manuscript.

### Appendices

#### Appendix 1 – Protocol for DNA Extraction

Each LBC sample was washed and re-suspended in 500μl 10 mM Tris pH 7.4. For every 22 samples, one positive control of 250μl Plasmid 51 (Deutsches Krebsforschungszentrum, Heidelberg) and one negative control of 250μl H20 were created. 50μl of recombinant proteinase K was added to a 250μl cell suspension from each sample and incubated at 56°C overnight with shaking. Samples were then incubated at 90°C for 10min, placed in racks (previously chilled to -20°C) in a 4°C fridge for 10min, centrifuged at 13 000 r.p.m, 4°C, for 10 min and the supernatant transferred into a 96-well plate.

#### Appendix 2 – Protocol for GP5+/6+ PCR ELISA

Samples were processed in batches of 22 and a two-tier method applied: (i) an initial PCR-ELISA with a cocktail of HR type-specific probes (ii) A second PCR-ELISA using the individual HPV 15 probe. PCR cycling conditions were 94°C – 4mins, (94°C – 30s, 40°C – 90s, 72°C – 60s; x 40), 72°C – 4mins, 15°C – hold). Positive (Plasmid) and negative (water) DNA extraction, PCR and ELISA controls were included for every 22 samples. The final negative extraction control in each 96-well plate serves as the background reading for which all the other results are compared. A positive result is equivalent to three times background.

#### Appendix 3 – Protocol for Linear E7 PCR

PCR was performed on extracted DNA for each sample including positive and negative controls. 5μl of DNA was added to 20 μl of PCR reagents (2.5μl 10x Invitrogen® buffer, 2.5μl 2mM dNTPs, 51 linear E7 primer 10μM 2.5μl, Invitrogen® Taq 1U 0.125μl, H2O 12.375μl). PCR cycling conditions were 95°C-15mins (94°C-30s, 62°C-30s, 72°C-3 min) 72°C-7 min, 4°C-hold).

#### Appendix 4 – Protocol for Nested E7 PCR

5μl of DNA was added to 20 μl of PCR reagents (2.5μl 10x Invitrogen® buffer, 2.5μl 2mM dNTPs, forward primer 5μM 2.5μl, reverse primer 5μM 2.5μl, MgCl_2_ 25μM 2.5μl, Invitrogen® Taq 1U 0.1μl, H2O 7.4μl). PCR cycling conditions were 95°C-15mins (94°C-30s, 58°C-30s, 72°C-30s) 72°C-5 minutes, 4°C-hold). Electrophoresis was performed on E7 PCR products. Results were photographed and compared to a 100bp DNA Ladder to determine positive or negative.

#### Appendix 5 – Protocol for E7 PCR using Hotstar Taq

5μl of DNA was added to 20 μl of PCR reagents (2.5μl hotstar buffer, 2.5μl 2mM dNTPs, forward primer 5μM 2.5μl, reverse primer 5μM 2.5μl, Hotstar Taq 1U 0.125μl, H2O 9.875μl). PCR cycling conditions were 95°C-15mins (94°C – 30s, 58°C-30s, 72°C-30s) 72°C-5 minutes, 4°C – hold). Electrophoresis was then performed on E7 PCR products. Results were photographed and compared to a 100bp DNA Ladder and results are then given as positive (1) or negative (0) and entered into an Excel worksheet.

